# Retinal implantation of electronic vision prostheses to treat retinitis pigmentosa: A systematic review

**DOI:** 10.1101/2020.11.30.20234476

**Authors:** Luke E. Hallum, Steven C. Dakin

**Affiliations:** Department of Mechanical Engineering, University of Auckland; School of Optometry and Vision Science, University of Auckland

## Abstract

**Purpose:** Retinitis pigmentosa is an hereditary disease causing photoreceptor degeneration and permanent vision loss. Retinal implantation of a stimulating electrode array is a new treatment for retinitis pigmentosa, but quantification of its efficacy is the subject of ongoing work. This review evaluates vision-related outcomes resulting from retinal implantation in participants with retinitis pigmentosa.

**Methods:** We searched MEDLINE and Embase for journal articles published since 1 January 2015. We selected articles describing studies of implanted participants that reported post-implantation measurement of vision. We extracted study information including design, participants’ residual vision, comparators, and assessed outcomes. To assess risk of bias, we used signalling questions and a target trial.

**Results:** Our search returned 425 abstracts. We reviewed the full text of 34 articles. We judged all studies to be at high risk of bias due to study design or experimental conduct. Regarding design, studies lacked the measures that typical clinical trials take to protect against bias (e.g., control groups and masking). Regarding experimental conduct, outcome measures were rarely comparable before and after implantation, and psychophysical methods were prone to bias (subjective, not forced-choice, methods). The most common comparison found was between post-implantation visual function with the device powered off versus on. This comparison is at high risk of bias.

**Conclusions:** There is a need for high-quality evidence of efficacy of retinal implantation to treat retinitis pigmentosa.

**Translational Relevance:** For patients and clinicians to make informed choices about retinitis pigmentosa treatment, visual function restored by retinal implantation must be properly quantified and reported.

## Introduction

Retinitis pigmentosa (RP) is a group of hereditary diseases causing photoreceptor degeneration^1,2^. Vision loss due to RP is progressive, and can result in total and permanent blindness. RP is a leading cause of blindness, afflicting 1 in 4000 people^3,4^.

In RP, the inner layers of the retina -- comprising ganglion, bipolar, and amacrine cells -- appear less vulnerable to degeneration than the outer layers, which comprise rod and cone photoreceptors^5^. Because RP predominantly affects outer layers of the retina, it has been proposed that retinal implants -- which attempt to replace the function of photoreceptors by electrically stimulating the retina’s more viable inner layers -- may be an effective treatment for RP (reviewed by Weiland and colleagues^6^). These vision prostheses typically comprise an electrode array that is surgically implanted either epiretinally (at the vitreoretinal interface), subretinally (between the photoreceptor layer and choroid), or suprachoroidally. That array is then coupled to an imaging light sensor device, such as a head-mounted camera, through a wired or wireless interface.

A recent review by Humayun and colleagues^55^ describes the development of retinal implants and, concomitant with this, the development of new methods for vision assessment in implantees. These new methods began with intra-operative detection of single phosphenes, and, as we discuss below, have recently expanded to include more conventional psychophysical tests (e.g., localization, spatial vision, and motion perception). As discussed by Humayun and colleagues, vision tests in implantees have generally not demonstrated measurable outcomes using conventional clinical approaches (e.g., letter charts). Therefore, the field’s pioneers were required to develop novel methods to attempt to measure clinical outcomes, running alongside technological developments, for the specific purpose of determining what a retinal prosthesis could do for a patient. The absence of measurable outcomes using conventional clinical tests need not imply that implantation confers no benefit^55^. To illustrate, patients with ultra-low vision, which is usually qualified (not quantified) in clinical terms such as “hand motion”, “light projection”, or “light perception”, do not exhibit measurable outcomes on conventional clinical tests, but nonetheless there are activities of daily living that appear to benefit from ultra-low vision (discussed by Humayun and colleagues^55, 60^). These are important considerations when evaluating vision-related outcomes of retinal implantation, and ought to be considered alongside our analysis, presented below, of the studies we discovered through systematic review. We acknowledge the methodological difficulty of testing a retinal prosthesis. At this stage of device development, our robust analysis is important for several reasons: it helps patients and clinicians make informed treatment decisions; it helps guide device design; and, it improves the specificity of retinal implant candidacy criteria.

Four retinal implant devices have been approved for use in either North America or the European Economic Area^7,8^, and there exist several pre-clinical experimental devices^9,10^. While these devices have received considerable attention in the media, quantification of their efficacy is the subject of ongoing work. In order for patients, clinicians, and funders to make informed decisions about experimental treatments for RP, it is vital that the efficacy of devices is properly quantified and reported. For patients, the primary outcome of interest will be the level of visual function that is restored following implantation. We therefore conducted a systematic review to evaluate vision-related outcomes resulting from retinal implantation of any device in participants with outer retinal degeneration. At the outset, we were primarily interested in the comparison of quantitative measures of vision made before and after implantation.

## Methods

### Data Sources and Study Selection

We searched MEDLINE and Embase for titles or abstracts containing any of our search strings. Search strings were all pairwise combinations of terms drawn from two sets. The first term was drawn from the following set: `retina`,`retinal`,`epiretina`, `epiretinal`, subretina, `subretinal`, `transretina`, `transretinal`. The second term was drawn from the following set: `implant`, `implantation`, `prosthesis`, `prosthetic`. An example search string is `retinal implantation`. We limited the search to journal articles published between 1 January 2015 and 11 October 2019 (date of last search: 14 October 2019). We screened abstracts resulting from the search to determine papers of primary interest. We then screened the full-text articles of these publications.

### Study Eligibility Criteria

We included studies of participants who (a) had undergone retinal implantation to treat outer retina degeneration and (b) had received a post-implantation assessment of vision. Assessments included both “functional vision”, as well as “visual function”; the former required participants to perform “real-world” visual tasks (e.g., to recognize household objects), and the latter required participants to perform psychophysical tasks involving constrained visual stimuli (e.g., to detect sine-wave gratings). We excluded “phosphene” studies (e.g., a study by Shivdasani and colleagues^11^) which, as opposed to presenting visual stimuli to implantees, electrically stimulated participants’ retinas with an acutely or chronically implanted electrode or electrode array. We included studies of any design, involving any device, including devices approved by the US Food and Drug Administration or an equivalent regulatory body, as well as experimental studies of devices yet to be approved.

### Data Extraction and Quality Assessment

We extracted data from each study using a custom data extraction form (Appendix A) modelled on a recent systematic review and meta-analysis of cochlear implantation by Gaylor and colleagues^12^. To guide our assessment of risk of bias, we used the signalling questions^13^ of Gaylor and colleagues^12^; in general, signalling questions are used to extract information about features of a study that are relevant to its risk of bias^13^. After our initial scoping of the review, we decided to add the following signalling questions:

- For interventional studies, were outcome assessment methods comparable before and after implantation?
- Were outcome assessment methods comparable with the device powered off and on?
- Do psychophysical methods appear sound (e.g., were forced-choice methods used where appropriate)?

To guide our assessment of risk of bias, we also referred to the Cochrane Handbook of Systematic Reviews of Interventions, specifically chapters 8 and 25^13,14^. To guide our reporting, we employed methods described in the PRISMA (Preferred Reporting Items for Systematic Reviews and Meta-Analyses) statement^15^.

## Results

Our searches returned 425 articles (including duplicates), of which 391 were excluded at the abstract level. We examined the remaining 34 full-text articles that met our predefined inclusion criteria. The selection process is illustrated in Figure 1. Seven devices were covered in the 34 articles considered (Table 1): three are epiretinal, two are subretinal, and two are suprachoroidal. We judged that all reviewed studies were at a high risk of bias^13,14^ for one or more of the following reasons: clinical trials were uncontrolled, neither participants nor outcome assessors were masked to experimental variables, and there were differences between pre- and post-implantation outcome assessment.

**Table 1:**
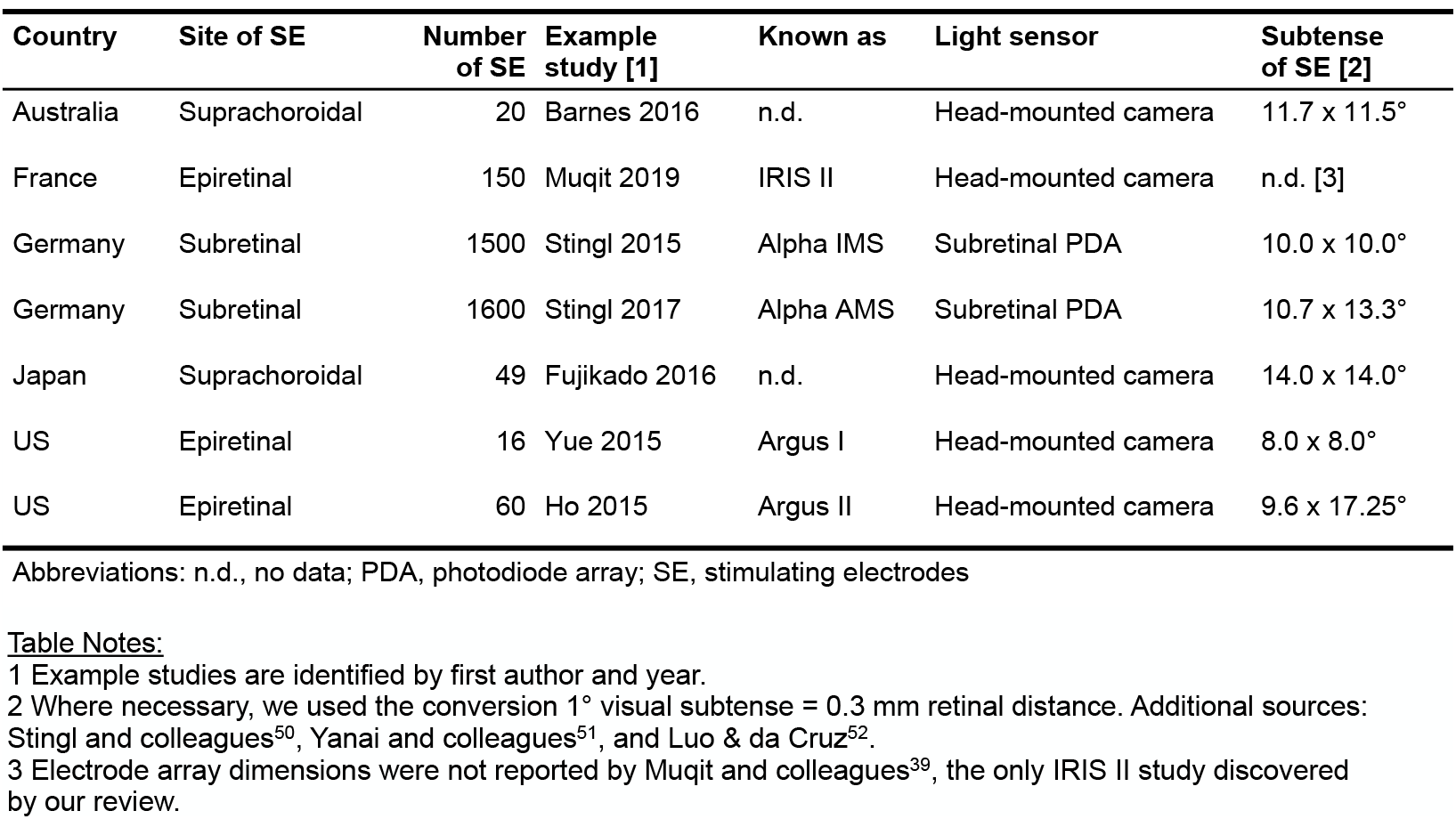
Retinal prostheses.

**Figure 1:**
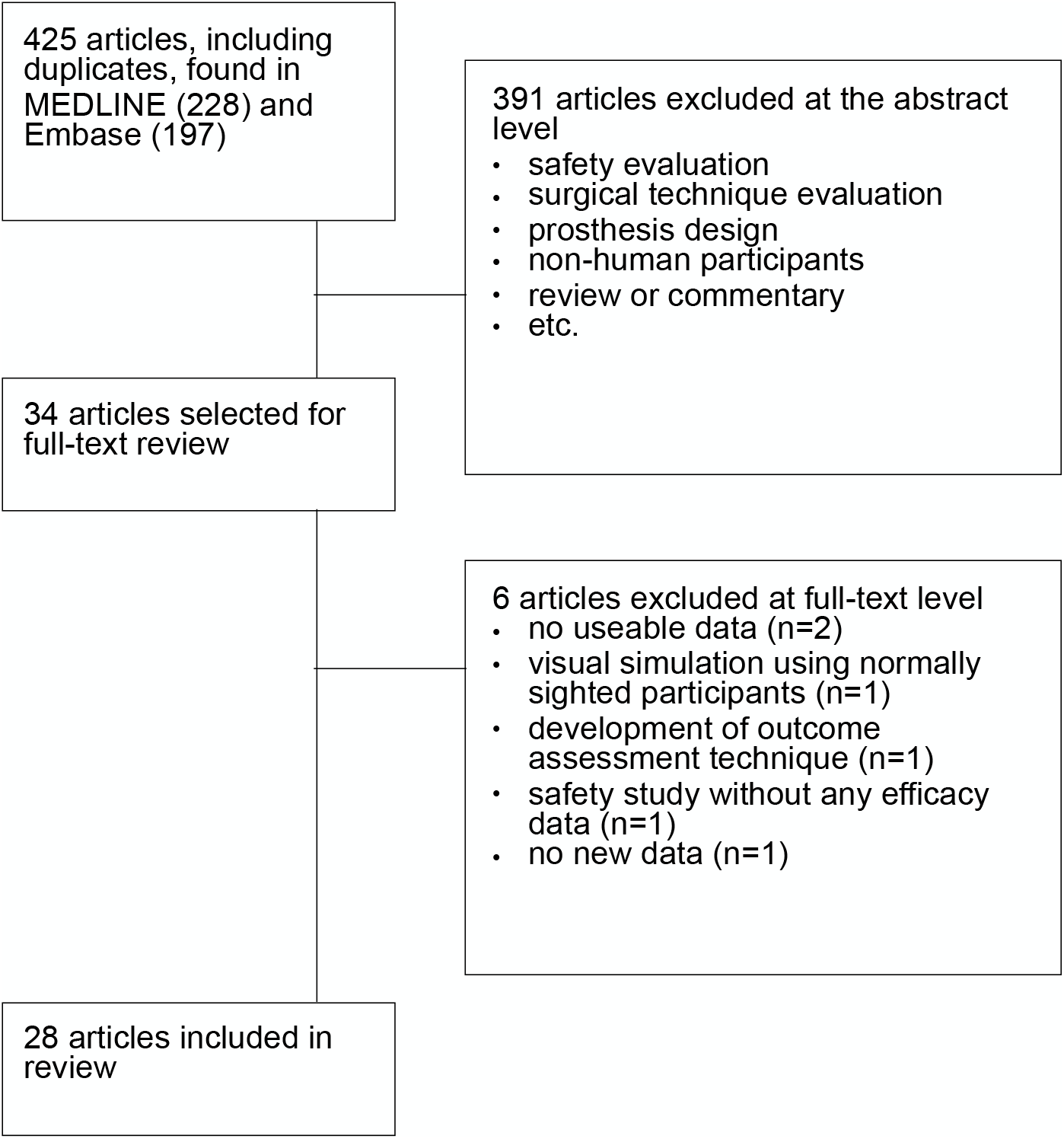
Flow diagram of the study selection process.

Our review revealed that the studies used a wide range of methods to asses visual outcomes, both before and after implantation. Straightforward interpretation of results was possible when studies used the more robust experimental methods typical of visual psychophysics. Such studies measured light perception, spatial vision, and motion perception (Table 2 and citations below). Many studies measured outcomes that were harder to interpret, some of which may have involved non-visual sensory cues. These outcomes were often associated with activities of daily living (e.g., sorting black, white, and grey socks), and therefore may ultimately play an important role in understanding the practical efficacy of retinal implantation.

**Table 2:**
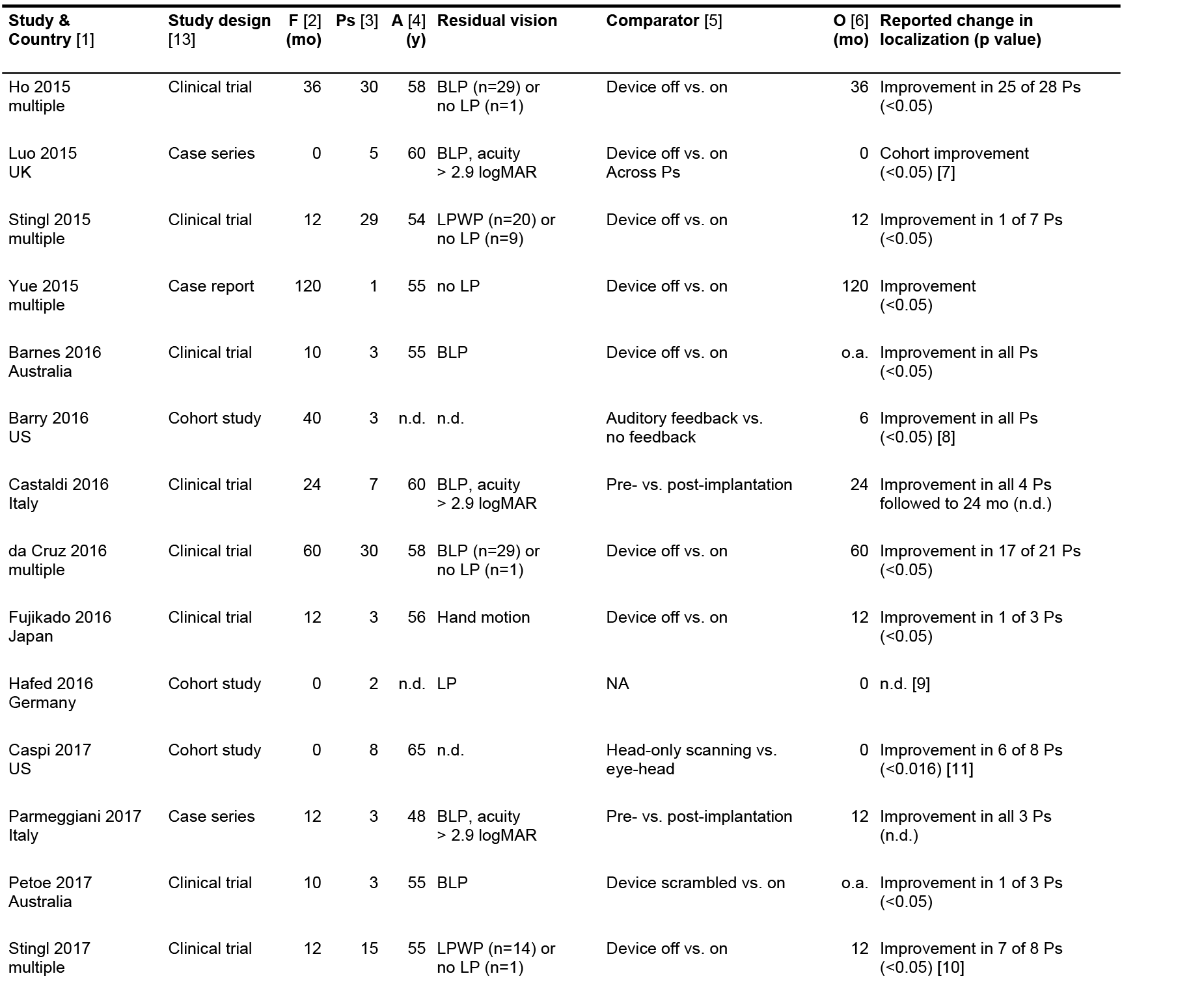
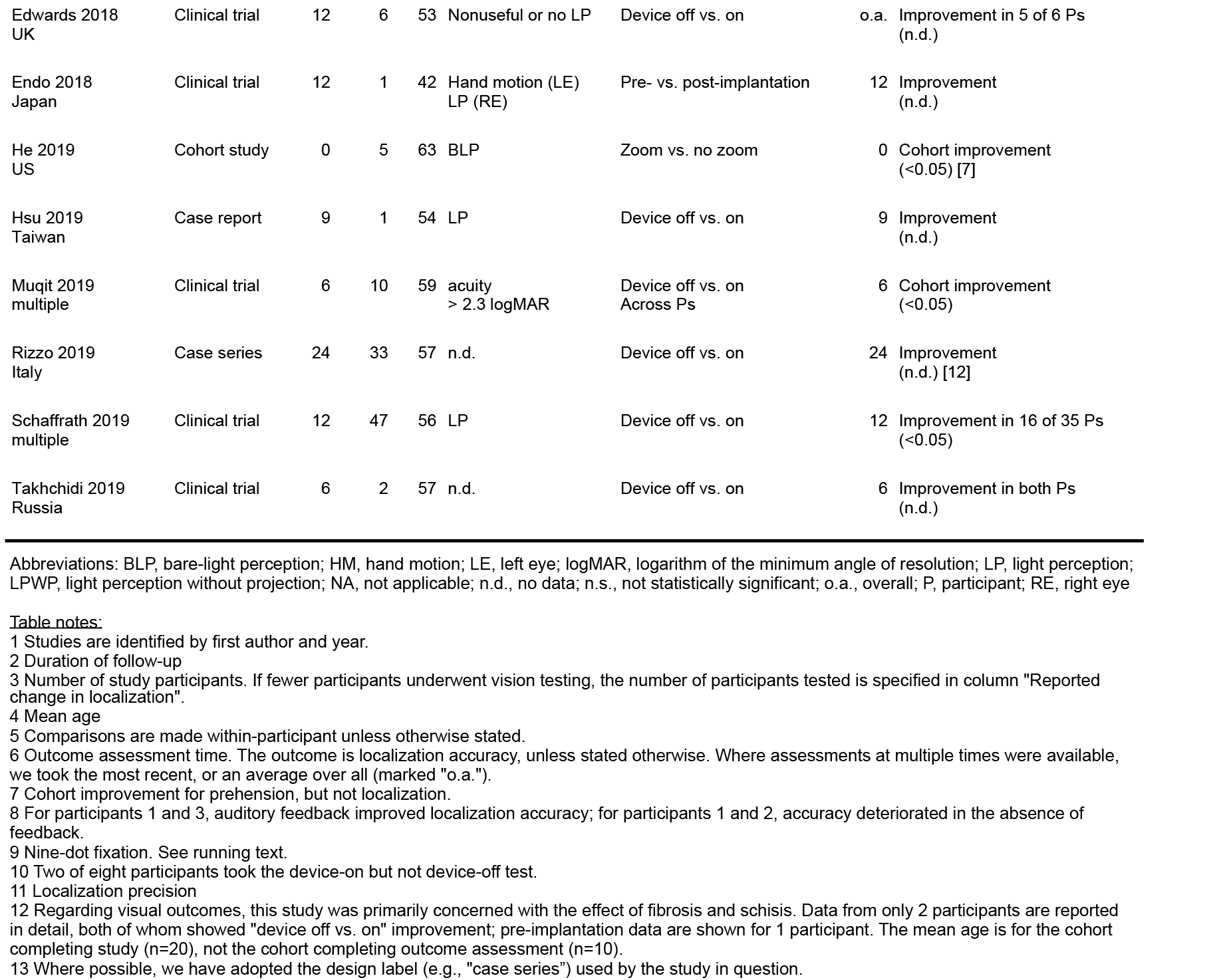
Localization-related outcomes of retinal implantation.

### Light perception

Our review discovered 22 studies that assessed the implanted participant’s ability to localize a light source (Table 2). This outcome was typically assessed using one of two test types. The first was a four-alternative forced-choice test using a high-contrast wedge stimulus appearing at one of four locations around a central fixation point^16^. Stimuli were typically large, e.g., subtending 20 degrees of visual angle, and were presented for long durations (e.g., the study by Barnes and colleagues^17^). The second test type involved patients indicating the location of a large (diameter approximately equal to 10 degrees) high-contrast square or circle appearing at a random location on a touch screen^18^. Stimuli persisted on screen until the participant’s touch landed (e.g., see da Cruz and colleagues^19^). Studies by Luo and colleagues^20^ and He and colleagues^21^ used a variation on the second test type by requiring participants to localize and reach to a high-contrast physical object (plastic blocks measuring 3.1×3.1×5.7cm, or drinking cups). Rather than using prehension or pointing, Hafed and colleagues^22^ measured the eye movements of two implanted participants. While their depiction of one participant’s pattern of fixation on a nine-dot calibration pattern (dot diameter = 2.4 deg) provides a qualitative assessment of localization accuracy, this accuracy was not quantified.

We discovered only three studies reporting localization accuracy both before and after implantation^23,24,25^. Accuracy refers to a participant’s ability to point to a target in an unbiased fashion (i.e., pointing may be variable, but, on average, pointing is to the correct location, and not, e.g., slightly to the left). This is in contrast to measures of precision, which quantify the reliability of a participant’s pointing (e.g., a participant always points slightly to the left, but this behaviour is consistent). These three studies^23,24,25^, following participants for up to 24 months, all reported pre- versus post-implantation vision improvement in all participants (Table 2). Most discovered studies compared localization accuracy when the device was powered off and compared that to accuracy when the device was on. Many of these studies were interventional (i.e., clinical trials) and followed participants (in one case, for 10 years), but others were observational (i.e., cohort studies). Most of these studies reported “off versus on” vision improvements in a majority of participants (Table 2). One clinical trial compared localization accuracy with the device on to accuracy after scrambling the phosphene map^10^; “normal versus scrambled” vision improvement was seen in only one (of three) participants. One cohort study compared localization accuracy with and without auditory feedback^26^; in all three participants the presence of feedback either increased accuracy, or its absence caused accuracy to decrease. Another cohort study integrated both eye- and head movements into phosphene activation^27^; localization precision was increased in six of eight participants when comparing eye-and-head to head-only scanning.

Several studies of localization accuracy requiring participants to point to targets (e.g., Hsu and colleagues^28^) did not report whether feedback was provided. This omission complicates the interpretation of results; without feedback, low accuracy may reflect participant bias as opposed to inaccurate light localization (as is the case with all psychophysical estimation tasks).

Participants can use feedback to adjust for any inherent bias^26^. Indeed, studies by Barry & Dagnelie^26^ and Caspi and colleagues^27^ are the only ones that explicitly addressed this complication; the latter (which reported precision, not accuracy) notes that pointing accuracy “should be a function of the position of electrodes on the retina, inherent point bias, and any differences in eye and [head-mounted] camera positions not taken into account.” It is unclear how to interpret many of the reported localization accuracies; if feedback was absent, does inaccuracy in performance indicate inherent bias, or inaccurate localization?

### Spatial Vision and Motion Perception

Our review discovered 10 studies that assessed implanted participants’ vision using sine- and square-wave grating stimuli^7,8,10,17,19,23,29,30,31,32^. This outcome was typically assessed using a two- or four-alternative forced-choice test, presenting a grating to the participant who was required to report its orientation. In all cases, stimulus duration was long, enabling scanning of stimuli with the head-mounted camera or implanted photodiode array: Ho and colleagues^29^ reported a grating stimulus duration of 5 seconds, presumably the same duration used by da Cruz and colleagues^19^; Stingl, Edwards, and colleagues^7,30,31^ all appear to have used “no strict time limits… participants were encouraged to provide prompt answers”^7^; Barnes, Petoe, and colleagues^10,17^ allowed participants to scan indefinitely before responding; and, Castaldi and colleagues^23^ used grating stimuli presented for a fixed duration of 1s. All studies measured each participant’s performance with the implanted device powered off, and compared that to device-on performance. However, none of the clinical trials compared grating acuity before implantation to grating acuity after implantation. Our review discovered reasonable consistency of experimental design across these studies (e.g., clinical trials with outcome assessment at multiple follow-ups, and the use of forced-choice methods). However the outcomes reported varied widely (e.g., one study reported spatial frequency discrimination threshold in units of cycles per degree, while another reported the percentage of their cohort with grating acuity better than 2.9 logMAR). All grating studies reported measurable grating acuity in implantees. Studies by Ho and colleagues^29^ and da Cruz and colleagues^19^ reported nine of 27 (33%; three-year follow-up) and eight of 21 (38%; five-year follow-up) participants, respectively, with grating acuity better than 2.9 logMAR. Petoe and colleagues^10^ (see also Barnes and colleagues^17^) reported grating acuity of 0.124 cycles per degree in one participant (in this participant, device-off discrimination threshold was unmeasurably low, while device-on thresholds were unmeasurably low in the cohort’s remaining two participants); Stingl, Edwards, and colleagues reported grating acuities upto 3.3 cycles per degree^7,30,31^; Castaldi and colleagues^23^ reported cohort performance on a grating detection task at rates well above chance; and Takhchidi and colleagues^32^ reported grating acuity less than 2.9 logMAR in both implanted study participants.

Taken together, the outcomes of the above-mentioned grating studies are encouraging. However, the interpretation of these results is not straightforward; it is difficult to separate the effect on performance of the following two factors: first, spatial vision, that is, perception arising from retinal patterns of activation; and, second, the combined effects of light perception and scanning. Indeed, when gratings are presented for long durations, participants are able to scan the stimulus, and a scanning participant can use integrated light perception to discriminate gratings of different orientations. We illustrate this concept in Figure 2. We are not suggesting that light perception and scanning exclusively account for the outcomes of the above-mentioned grating studies. Rather, we emphasize that these outcomes conflate spatial vision and light perception to an unknown extent. In other words, if implanted participants are allowed to scan grating stimuli, then caution must be exercised when using the outcomes of those experiments to draw conclusions about the restoration of spatial vision in the absence of any scanning. While the stimulus duration reported by Castaldi and colleagues^23^ was relatively short (1 second), their detection task involved “stationary, low spatial frequency” gratings which participants may have learned to scan. Grating studies by Stingl, Edwards, and colleagues^7,30,31^ involved only a small number of trials and/or reported each participant’s “best achieved” grating acuity^30,31^, leaving the results vulnerable to false-positive findings.

**Figure 2:**
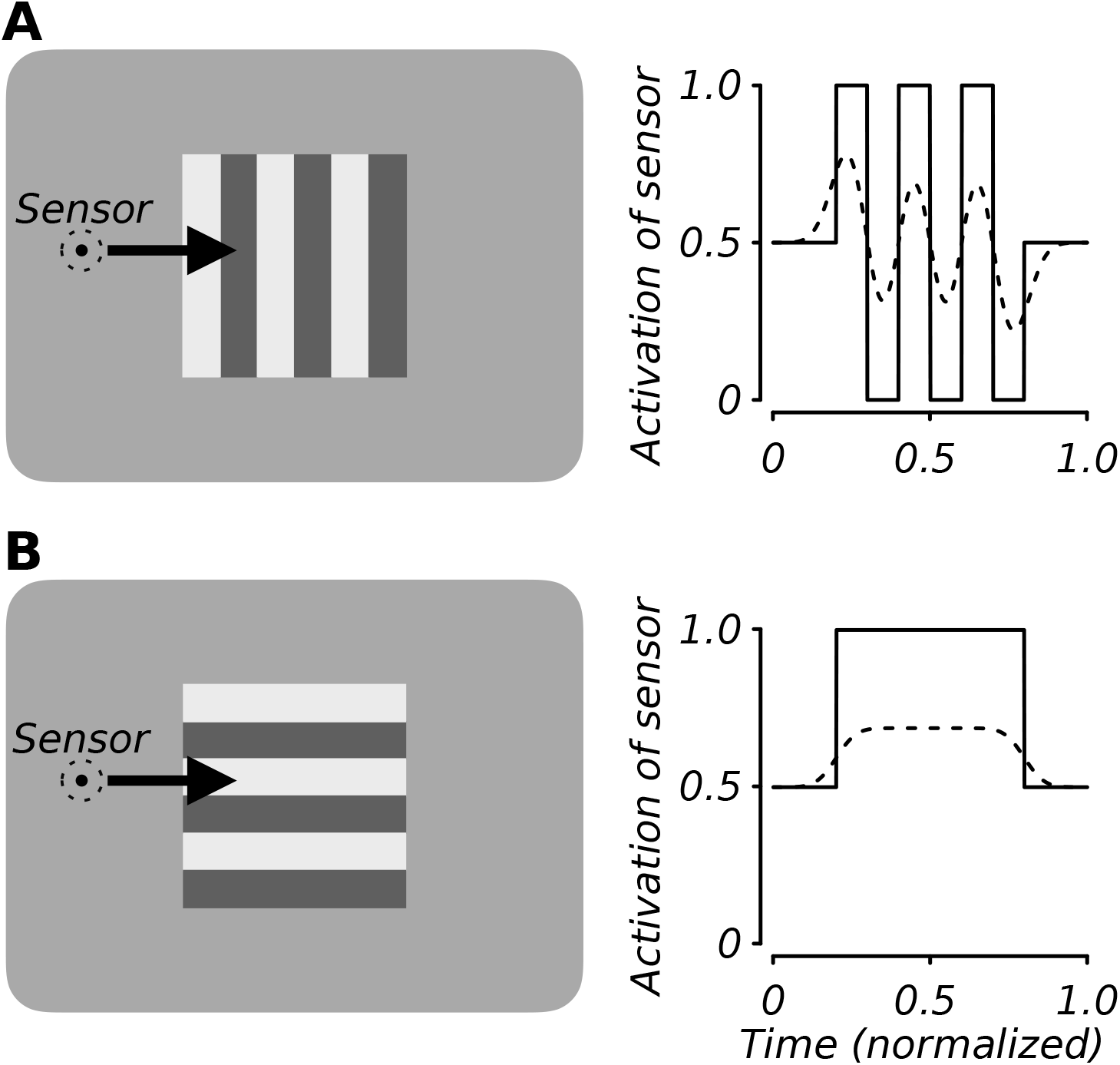
A non-imaging light sensor (e.g., a photodiode) can be used to discriminate oriented grating stimuli. (**A**) An example vertical grating appears on a computer display (left). A sensor scans the display from left to right (arrow). The sensor’s activation versus time is illustrated in the right panel (time 0 corresponds to the onset of scanning, and 1 corresponds to its cessation). If the sensor’s (i.e., photodiode’s) aperture is large, the efficacy of scanning is reduced. This trade-off is illustrated by the dashed circle, representing a larger aperture, superimposed on the stimulus; for a larger aperture, the modulation of the output is reduced (dashed activation of sensor at right). For very large apertures, the modulation of the output is reduced to zero. (**B**) As in (A), but the grating is horizontal. The different temporal patterns of activation can be used to discriminate vertical gratings from horizontal. A scanning strategy may be employed by an implanted participant; the participant’s ability to localize light may then be mistaken for spatial vision.

The image processing implemented by a retinal prosthesis limits the efficacy of scanning; gratings beyond the passband of a regional averaging filter (e.g., see simulations by Chen, Hallum and colleagues^57,58^), or a similar low-pass filter, are greatly attenuated, and therefore no amount of scanning will enable the participant to discriminate these relatively high-spatial-frequency gratings. However, scanning relatively low-spatial-frequency gratings (i.e., within the passband) is an effective strategy. The participant tested by Barnes and colleagues^17^ scanned relatively low-spatial-frequency gratings for long periods (on average, 44.5 s per trial) to achieve grating acuities of between 0.09 and 0.124 cyc/deg, that is, acuities within the passband of their device. In Figure 2, we illustrate the effect of regional averaging, or a similar low-pass filter. As the averaged region increases in size (dashed circle superimposed on the stimulus in Figure 2), the modulation of the photodiode output is reduced. In other words, the efficacy of scanning is reduced as the averaged region increases. We also note that there is a potentially useful comparison to be made between the functional role of involuntary fixational eye movements and voluntary scanning, which contribute to normal and prosthetic vision, respectively.

Our review discovered four studies that used either drifting gratings or dot stimuli to test participants’ motion perception^7,23,30,31^. These are relatively strong tests of motion perception. Participants performed no better than chance in two of these studies^7,23^. Stingl and colleagues^30,31^ reported that a minority of participants passed a dot motion test at the most recent follow-up, however these experiments were underpowered. Several studies of participants’ motion perception used a drifting, high-contrast bar as the stimulus, requiring participants to estimate its direction of motion^19,29,33^. Albeit the outcomes of these moving-bar studies are encouraging, the interpretation of results is not straightforward; analogous to the above-described grating studies, in moving-bar studies it is difficult to separate the effect on performance of two factors: motion perception and light perception. To illustrate, consider an implanted participant fixating the right side of the stimulus monitor; this participant could use the time of arrival, there, of light to estimate direction of motion at rates above chance. We illustrate this concept in more detail in Figure 3. We are not suggesting that light perception exclusively accounts for the outcomes of the above-mentioned moving-bar studies. Rather, we emphasize that these outcomes conflate motion perception and light perception to an unknown extent. We note that implanted participants may have difficulty stabilizing fixation at a point on the stimulus monitor (in moving-bar studies, participants were “instructed to maintain eye and camera (head) fixation on the center of a 19-inch touchscreen monitor”^36^); this instability would add uncertainty to timing information, rendering it less useful in the performance of the task. Uncertainty could also be added to timing information by introducing an interval, randomized across trials, between the beginning of a trial and the onset of bar motion. However, none of the studies we discovered reported the use of such an interval^19,29,33,36^. We also note that if an implanted participant adopted a strategy of fixating a peripheral location on the stimulus monitor (as we illustrate in Figure 3A), then to do so would be to go against the experimental instructions. If a participant fixated the center of the stimulus monitor (as instructed), then timing information would not be useful in performing the task.

**Figure 3:**
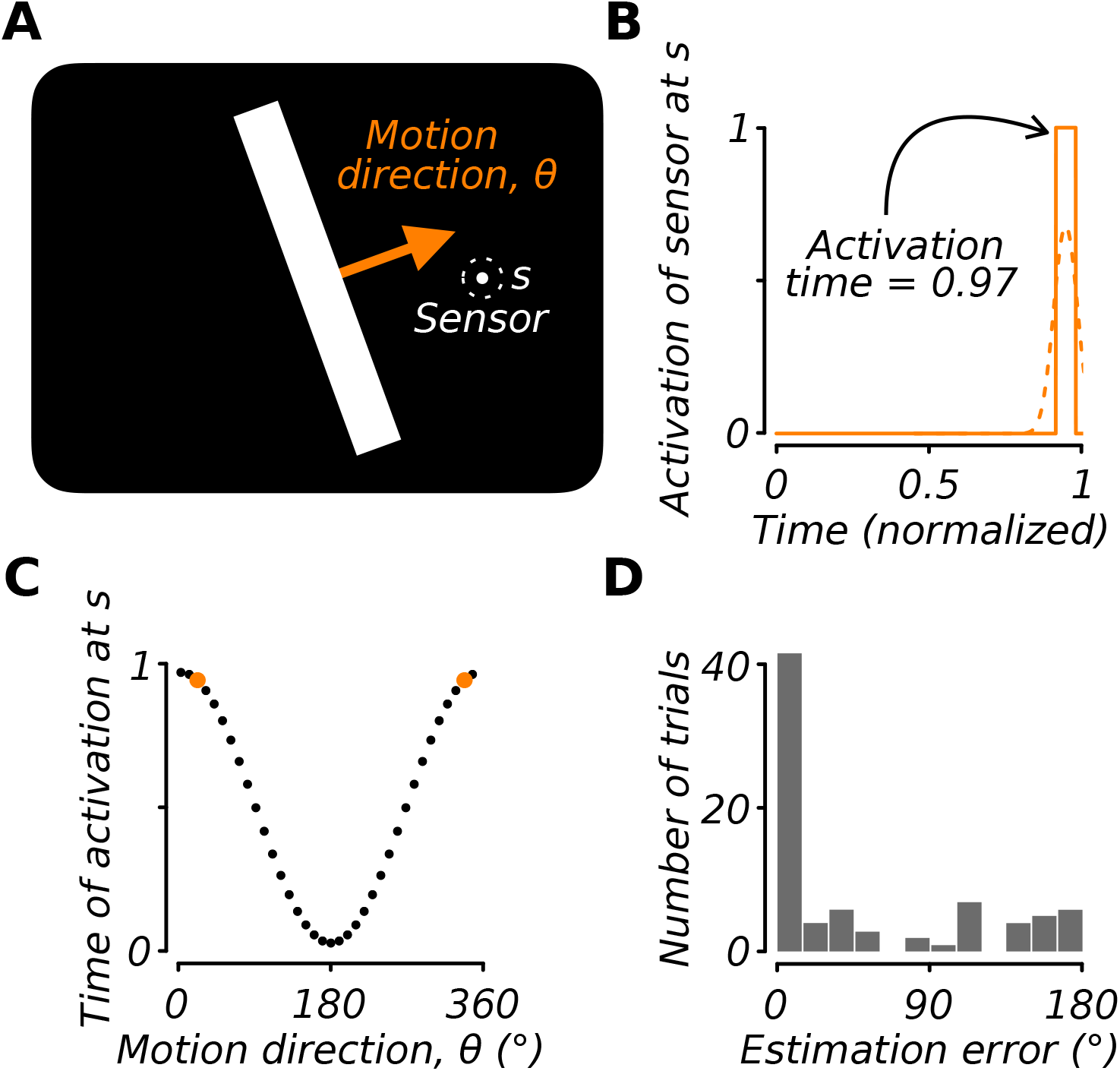
A non-imaging light sensor (e.g., a photodiode) can be used to discriminate motion direction of a high-contrast bar. (**A**) Example high-contrast bar moving across a computer display. The bar’s direction of motion (arrow) is 20 degrees and its speed is constant. A sensor is fixed at display location marked “s”. (**B**) The sensor responds to light when the bar traverses location “s”. The time of activation of the sensor is 0.97 (normalized), where 0 corresponds to the onset of the moving bar and 1 corresponds to its offset. If the sensor’s aperture is large (dashed circle in A), the time of maximum activation is unchanged (0.97), but the activation amplitude is reduced (dashed line). (**C**) The time of activation of the sensor at location “s” versus the direction of motion of the high-contrast bar. Here, we plot 36 directions equally spaced by 10 degrees. Each time of activation is consistent with two equally possible directions of motion. E.g., a time of activation = 0.97 is produced by either bar motion at 20 or 340 degrees (orange symbols). Time of activation can be used to estimate motion direction; given activation time = 0.97, a simple decoding strategy is probability matching^34,35^, i.e, to choose 20 or 340 degrees with equal probability. (**D**) Histogram showing estimation errors accumulated over 80 trials of a probability-matching simulation (motion direction was randomized across trials using a uniform distribution spanning 0 to 360 degrees). The majority of estimates were correct, i.e., within 15 degrees of the bar’s true direction of motion. This criterion (15 degrees) was used by Dorn and colleagues^36^ and many of the studies discovered by our review. In other words, a non-imaging light sensor combined with a simple decoding strategy passed the motion test without motion information. If an implanted participant employs a similar strategy, estimates of that participant’s motion perception will be biased. However, to adopt such a strategy, a participant would need to fixate a peripheral location of the stimulus monitor, going against the experimental instructions.

### Other Measures of Vision

Our review discovered that a wide range of other tests of vision are being used to assess the outcome of retinal implantation (several of which were questionnaire-based^7,19,29,33,40,41^). Many of these other tests attempted to quantify “real-world” vision and included the following:

- Landolt C optotype identification^17,30,31^
- “Door task”: patients attempted to walk to and touch a “simulated door”, that is, a large piece of contrasting felt on a wall^19,29,33^
- “Line task”: “patients followed a white line painted on black tiles”^9,19,29,33^
- Navigation in a controlled environment^37,38^
- Sock-sorting^38^
- Walking direction discrimination task^38^
- Tableware recognition^7,9,30,31^
- Analog clockface reading^7,30^
- Sensitivity to a full-field luminance increment^7,30,31^
- Temporal resolution^7^
- Picture recognition and Goldmann field testing^39^
- Letter acuity^30,54^
- Greyscale discrimination^30,31^

Tasks modelled on activities of daily living, such as those listed above, are potentially useful in understanding the practical efficacy of implantation. To illustrate, here we describe some outcomes relating to navigation (door and line tasks, navigation in a controlled environment, and walking direction discrimination)^9,19,29,33,37,38^ and object recognition (tableware recognition, clockface reading, letter acuity)^7,9,17,30,31^. da Cruz and colleagues^19^ reported improved performance on their door and line tasks at all post-implantation follow-ups to five years. Their reported door-task success rate at 5-year follow-up, averaged across a cohort of 20 participants, was approximately 50% (device on) versus approximately 20% (device off). Their reported line-task success rate (5-year follow-up, n=20) was approximately 70% (device on) versus approximately 20% (device off). However, line-following outcomes reported by Fujikado and colleagues^9^ were mixed. Two of their three participants showed improved performance (device off versus on), that is, less deviation from the walked line, at two of six and three of six follow-up sessions, respectively. However, in their third participant, the device’s power status had no effect on performance at any of the four follow-ups (all participants were followed for 12 months). Garcia and colleagues^37^ reported the performance of four implanted participants navigating a controlled environment, concluding that the implanted device did not provide sufficiently reliable visual information to improve navigation. On the first of two tasks (“path reproduction”), the implanted cohort (n=4) showed no improvement when navigating with the device on as compared to device-off navigation. However, on the second task (“path completion”), two of the implanted participants showed device-on versus off improvement comparable in magnitude to that of a control group (control participants were normally sighted, but wore goggles restricting field of view, and decreasing acuity to 1.6 logMAR). Dagnelie and colleagues^38^ reported the performance of 27 implanted participants navigating a controlled environment, or discriminating the walking direction of another person. On the former task (“sidewalk tracking”), the performance of approximately two-thirds of participants improved (fewer “out of bounds” deviations from the sidewalk) with the device on versus off; conversely, approximately one-third of participants performed better with the device off versus on. Overall, that cohort’s performance improved with the device on. On the latter “walking direction” two-alternative forced-choice task, the performance of 18 participants was significantly above chance (50% correct) with the device powered on. Only six participants performed significantly above chance with the device powered off.

Several of the studies we discovered reported outcomes concerning the recognition of objects^7,9,17,30,31^. Fujikado and colleagues^9^ tested participants’ ability to discriminate a bowl from chopsticks. Two of their three participants showed improved performance (device off versus on) at one of seven and five of six follow-up sessions, respectively. In their third participant, the device’s power status (on versus off) had no effect on performance at any of the five follow-ups. In a similar task, Edwards and colleagues^7^ reported that their five implanted participants were able to locate household objects (knife, fork, spoon, and plate) on a tabletop when the device was powered on, but not when the device was off. Two participants were able to name, as well as locate, objects in follow-up sessions 3 months after implantation. Naming was not possible with the device powered off. Similar outcomes were reported by the same group in a larger cohort^31^. Barnes and colleagues^17^ reported Landolt-C acuity in one implanted subject (of a cohort of three). They found a performance improvement (device off versus on) of 3.24 versus 2.65 logMAR. Stingl and colleagues^31^ also reported mixed acuity outcomes; they were able to measure Landolt-C acuities of 20/546 and 20/1111 (1.44 and 1.74 logMAR, respectively) in two of their 13 implanted participants.

Taken together, the outcomes of these studies involving tasks modelled on activities of daily living, are encouraging. However, a key question remains: To what extent did participants perform these tasks using scanning and light perception, and to what did they use spatial vision or motion perception? Data are consistent with the interpretation that the implant may afford participants improved light perception, and this improvement in turn affords improved performance on a diverse range of visual outcomes.

## Discussion

The studies discovered by our review indicate that retinal implantation of an electrode array to treat retinitis pigmentosa may be effective in restoring some degree of light perception. However, outcomes relating to spatial vision and motion perception, while encouraging, were open to more than one interpretation. In most discovered studies, neither participants nor assessors were masked to experimental conditions, and control groups were not used as comparators. In clinical trials, outcome assessment was rarely comparable before and after implantation. For these reasons, all discovered studies were judged to be at high risk of bias^13,14^. Our findings signal an urgent need for the development of objective protocols for vision assessment to be used at multiple sessions both before and after implantation.

We have applied robust analysis to studies discovered by our review. We acknowledge that high risk of bias is a likely finding given we reviewed several feasibility studies of devices in development, and we do not mean for our analysis to mitigate the interdisciplinary achievement represented by these human trials. However, at this early stage of device development, our robust analysis -- specifically, identifying risk of bias, and scope for improved study design – is important for two reasons. First, for patients and clinicians to make informed choices about retinitis pigmentosa treatment, vision restored by retinal implantation must be properly quantified and reported. Second, proper quantification and reporting could be used to guide device design, and to improve the specificity of retinal implant candidacy criteria. Improved design and candidacy criteria will help speed the development of proven, effective devices.

We focussed our analysis of studies using two key questions. First, is there evidence of retinal implants restoring light perception? This question is important; conclusive evidence that participants with no light perception before treatment can perceive light after treatment would help justify the implantation of a stimulating electrode. In low-vision patients, light perception is typically assessed in the clinic via qualitative methods. In experimental participants, light perception is typically assessed using quantitative, forced-choice methods and multiple trials; an observer either discriminates which of two temporal intervals contains a light increment, or which of two or four spatial locations contains a light increment. Second, is there evidence of retinal implants restoring spatial vision or motion perception? This question is centrally important; an affirmative answer would largely justify the implantation of multi-electrode arrays, as opposed to a single electrode. In a typical grating acuity assessment, an observer discriminates briefly presented spatial patterns defined by sine- or square-wave variations of luminance (“gratings”) that are different in spatial frequency and/or orientation. Gratings are, typically, briefly presented (<200 ms) to minimize the effect of eye movements on performance. Because the mean luminance of gratings does not vary as a function of spatial frequency and/or orientation, grating acuity assessment measures spatial vision^42^. Motion perception is a rigorous assessment of retinal implant efficacy as it requires both spatial and temporal analysis of visual information; implant recipients able to discriminate grating stimuli, or random dot stimuli, that drift in different directions but are otherwise identical, would also provide evidence to justify the implantation of multi-electrode arrays. By analogy, cochlear implant scientists, engineers, and clinicians have devoted much effort to demonstrating the clinical benefit of multi-channel devices as compared to single-channel devices (e.g., Tyler and colleagues^43^). Retinal implantation of multi-electrode arrays has yet to meet the same standards of quality of evidence. We note that the restoration of some spatial vision and motion perception is not the only justification for implanting a multi-electrode array; since the effects of RP can vary across the retina, implanting an array, as opposed to a single electrode, potentially increases the chance of contacting retina suitable for stimulation and, in turn, restoring some light perception.

Most of the studies discovered by our review measured a participant’s vision with the device powered off and compared that to performance measured with the device powered on. This comparison is problematic for several reasons. First, it does not assess the efficacy of retinal implantation as compared to no treatment, although it invites this interpretation. Indeed, retinal implantation of an electrode array may have a deleterious effect on residual vision. Second, the comparison is vulnerable to bias because in none of the studies we discovered were participants or outcome assessors masked to the power status of the device (power off versus on). A lack of masking is, for example, common in surgical trials and can bias outcomes^44^:

> “If participants are not blinded, knowledge of group assignment may affect their behaviour in the trial and their responses to subjective outcome measures. For example, a participant who is aware that he is not receiving active treatment may be less likely to comply with the trial protocol … Those aware that they are receiving or not receiving therapy are more likely to provide biased assessments of the effectiveness of the intervention -- most likely in opposite directions -- than blinded participants^45^. Similarly, blinded clinicians are much less likely to transfer their attitudes to participants or to provide differential treatment to the active and placebo groups than are unblinded clinicians^45^.”

In the context of retinal implantation, the lack of masking of participants to experimental variables may alter their behaviour in a direction that favours the device when powered on. The lack of masking of outcome assessors renders them more likely to provide differential interaction with participants contingent upon the power status of the device in a fashion that may favour the device when powered on. Unmasked studies wherein the outcomes assessed are subjective are especially vulnerable to bias^46^; all vision measurements are subjective, however forced-choice methods are criterion-free and, it stands to reason, less vulnerable to bias. We concede that masking participants is methodologically difficult where stimulation elicits clear, suprathreshold sensations. Nonetheless, participant masking deserves greater consideration; we discovered only one study (by Petoe and colleagues^10^) that discussed the potential consequences of a lack of masking on outcomes. That study reported altered behaviour in one of the three participants when the device was powered off. Indeed, those researchers designed additional experiments to attempt to control for “a perceived lack of participant engagement in tasks when using system off. We observed this situation in [participant] P2, corresponding to a significantly eccentric starting position and minimal head movement (i.e., small response offset), in spite of our instruction to attempt the task with residual vision.”

Our review discovered only three clinical trials that reported comparable vision assessments before and after implantation^23,24,25^. All three of these studies reported vision improvement after implantation. All other clinical trials described pre-implantation vision in only the qualitative terms of clinical assessment, not directly comparable to post-implantation measures (e.g., “In all 6 participants, this [pre-implantation vision] was recorded as vague nonlocalizing perception of light in the eye to be implanted”^7^). This, too, is problematic since without comparable outcome assessment before and after implantation, it is not possible to separate the conflated effects of the device and of residual vision on performance. Several studies implied that participant performance with the device powered off provides an estimate of residual vision. However, outcome assessment with the device off is potentially biased as discussed above, and therefore a poor estimate of residual vision. Furthermore, retinal implantation may have an overall deleterious effect on participants’ vision; the “device off versus on” comparison is insensitive to any deleterious effects. The best way to quantify the effect of residual vision on an implanted participant’s performance is to make comparable, quantitative measurements before and after implantation. Pre-implantation measurements would facilitate an answer to the central question: Is retinal implantation effective as compared to no treatment? At present, this is an open question.

The placebo effect -- benefit resulting solely from the administration of treatment -- may arise due to a variety of factors, including participant expectation of an effect^47^. The placebo effect is an important consideration in drug, surgical, and device trials alike^48^, especially where subjective outcomes are assessed. None of the clinical trials discovered by our review attempted to control for the placebo effect. Two studies, both of which made questionnaire-based vision assessments before and after implantation, discussed the placebo effect as a potential confound^40,41^. Both studies argued that, because their participants’ improved vision persisted throughout long follow-up periods, their results were “most likely not the result of placebo, which is known to deteriorate over time”^41^.

In reviewed studies, visual performance was variable, whether compared across participants within a single study of a particular device, or across participants drawn from studies of different devices. An understanding of the sources of this variability could be used to refine the design of devices and the criteria used to select candidates for implantation. At present, there are too few published data, and the outcomes assessed are too varied, to attempt to quantify factors predictive of visual outcomes of implantation. Indeed, there are likely many factors, including “genetic subtype of retinal disease, age at time of implant, age at time of total vision loss, apposition of the array and retinal surface, and many others”^8^. The identification of prognostic factors is an ongoing challenge to retinal implant engineers, scientists, and clinicians.

We used a “target trial” to guide our judgements of risk of bias of the uncontrolled clinical trials discovered by our review. A target trial is a hypothetical trial designed in such a way that it lacks any features that put assessed outcomes at risk of bias. The target trial need not necessarily be ethical, nor feasible^14^. This target trial may guide the design of future studies of retinal implantation -- or array implantation at other sites in the visual pathway -- and for that reason we describe its main features here. Our target trial enrolled a cohort of retinitis pigmentosa sufferers. Participants were randomly allocated to one of two groups; participants and outcome assessors were masked to this allocation. Prior to implantation, all subjects participated in the forced-choice experiments for the assessment of light perception, spatial vision, and motion perception, as described by Bach and colleagues^16,49^. The use of multiple pre-implantation follow-ups ensured that practice effects were at asymptote at the time of implantation. These assessments were made monocularly in each eye (with the fellow eye patched), as well as binocularly. At implantation, participants in one group received an active device, while those in the other group received a sham device. The only difference between the sham device and the active device was that, during the trial, the electrodes comprising the sham device carried no current (the sham device was designed to be activated at the conclusion of the trial so as not to dissuade participation). After implantation, all subjects participated in the same (monocular and binocular) forced-choice experiments for the assessment of light perception, spatial vision, and motion perception. The use of multiple post-implantation follow-ups enabled participants in the active group to learn to use the device, thus further emphasizing any between-group differences in outcomes. Combined with appropriate statistical analysis of repeated-measures data, this target trial facilitates two main comparisons: first, the comparison of vision outcomes between active and sham groups; second, the comparison of vision outcomes before and after implantation separately for each group. Thus, our target trial controls for residual vision, participant engagement, and the placebo effect. Systematic review with meta-analysis is the gold-standard for the evaluation of medical interventions; our target trial facilitates a future meta-analysis by using an assessment method, readily available and well described, that has been validated in low-vision populations^16,49^. An added benefit of assessing only three vision outcomes (light perception, spatial vision, and motion perception) is that false-positive findings are reduced (by contrast, some studies discovered by our review used many vision tests, in addition to collecting anecdotal reports of participants’ experiences). Our target trial would go a long way to providing strong evidence for the retinal implantation of an electrode array for treatment of retinitis pigmentosa. The provision of strong evidence, at present, is an ongoing challenge that lacks no urgency.

Since submitting this systematic review to *Translational Vision Science & Technology* (on 15th January, 2020), two noteworthy Perspectives have been published^56,59^. The first proposes guidelines for performing and reporting vision tests in participants implanted with a prosthetic device^56^. Several of these guidelines address issues (i.e., risks of bias) similar to those that we have raised above. For example, Bailey and colleagues (subsection Visual Acuity) clearly articulate the need for comparable pre- and post-implantation measurements when assessing visual acuity. The second Perspective articulates the pitfalls of attempting to test for vision by using methods design to measure the limits of a visual system that is assumed to function (normally or partially)^59^. Therein, Peli identifies problems with several tests currently in use to assess vision restoration, arguing that the field’s progress is contingent on the use of appropriate tests, free of bias, to guide the design of new and better devices.

## Data Availability

All relevant data are within the manuscript and its Supporting Information files.

## Acknowledgements

We thank Dr Shaun Cloherty and Prof. Eli Peli for comments on the manuscript. Thanks also to Prof. Peli for raising to our attention his recent presentation at the Annual Meeting of ARVO, “Testing for vision rather than testing vision” (Invest. Ophthalmol. Vis. Sci. 2019; 60(9): 2844).

## Appendix A

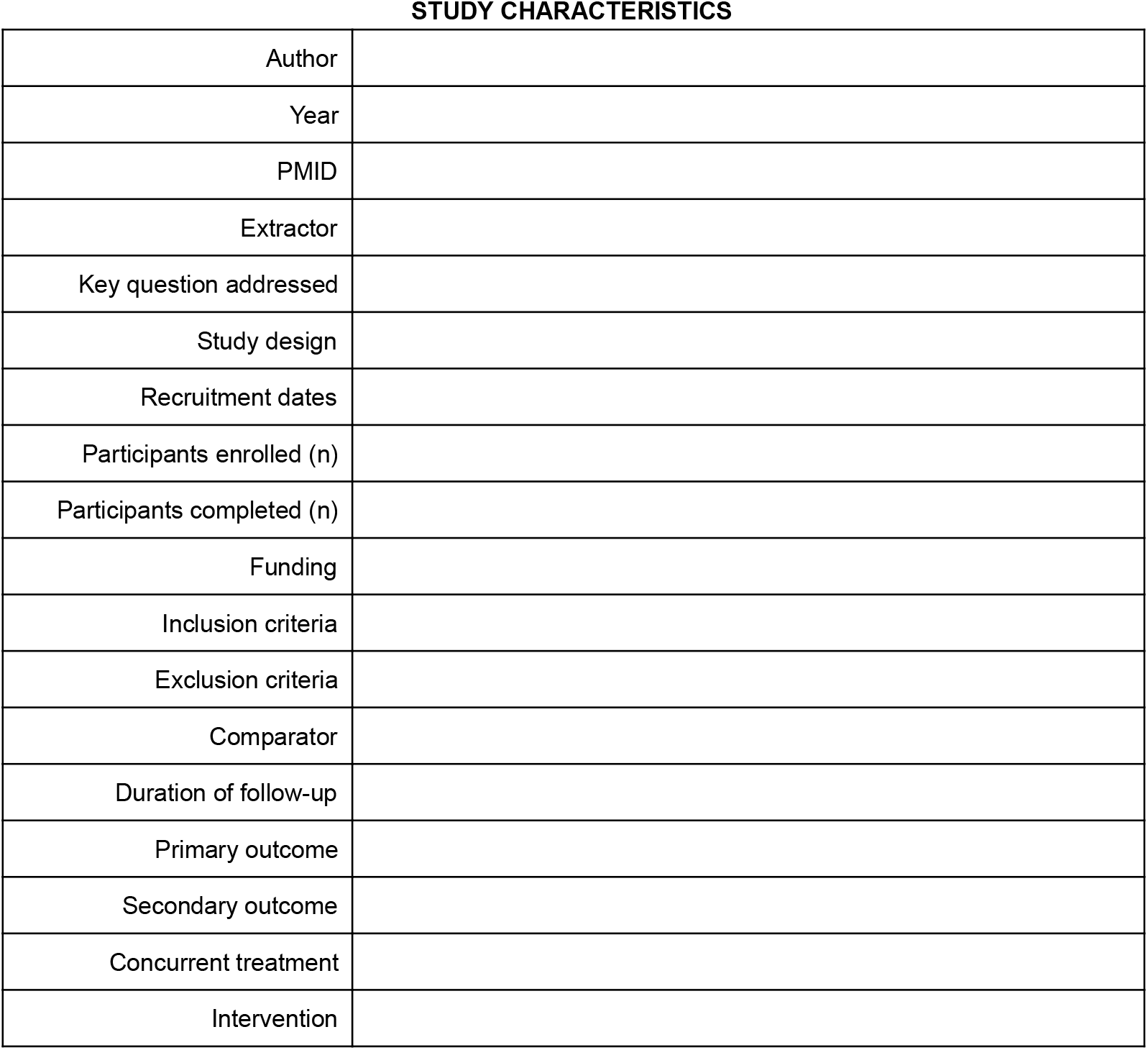
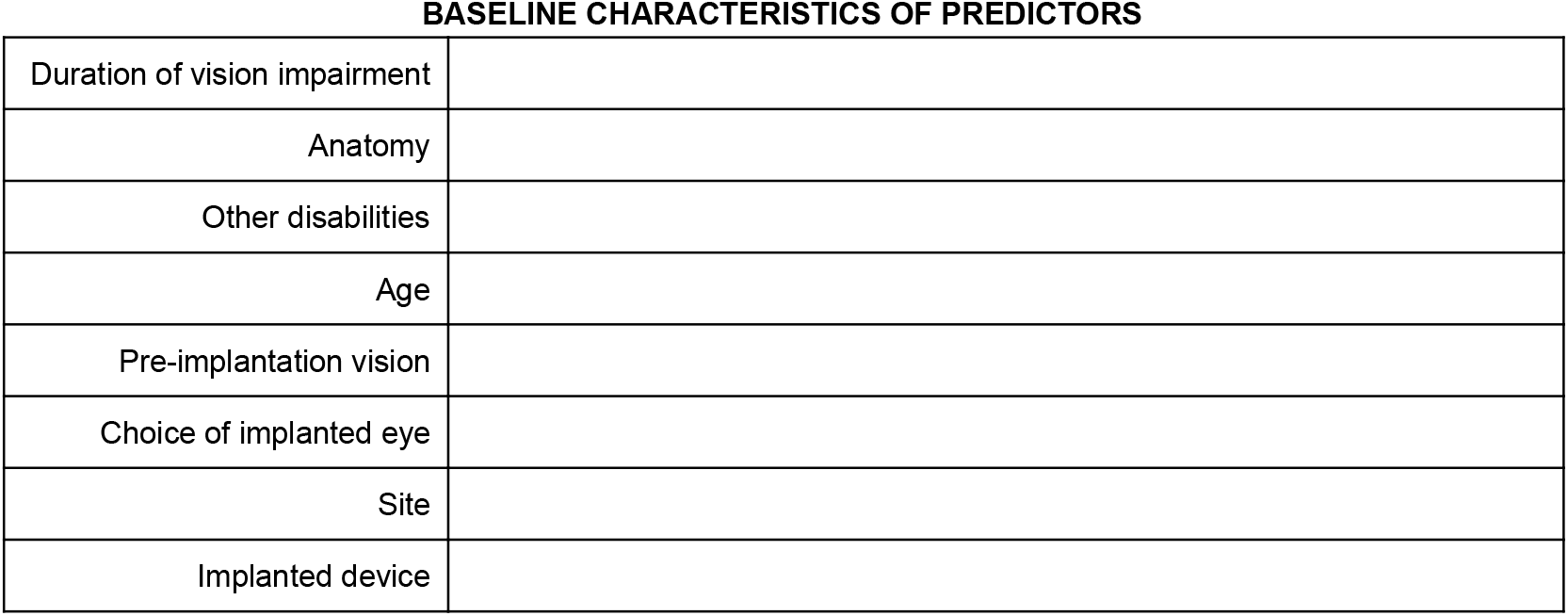
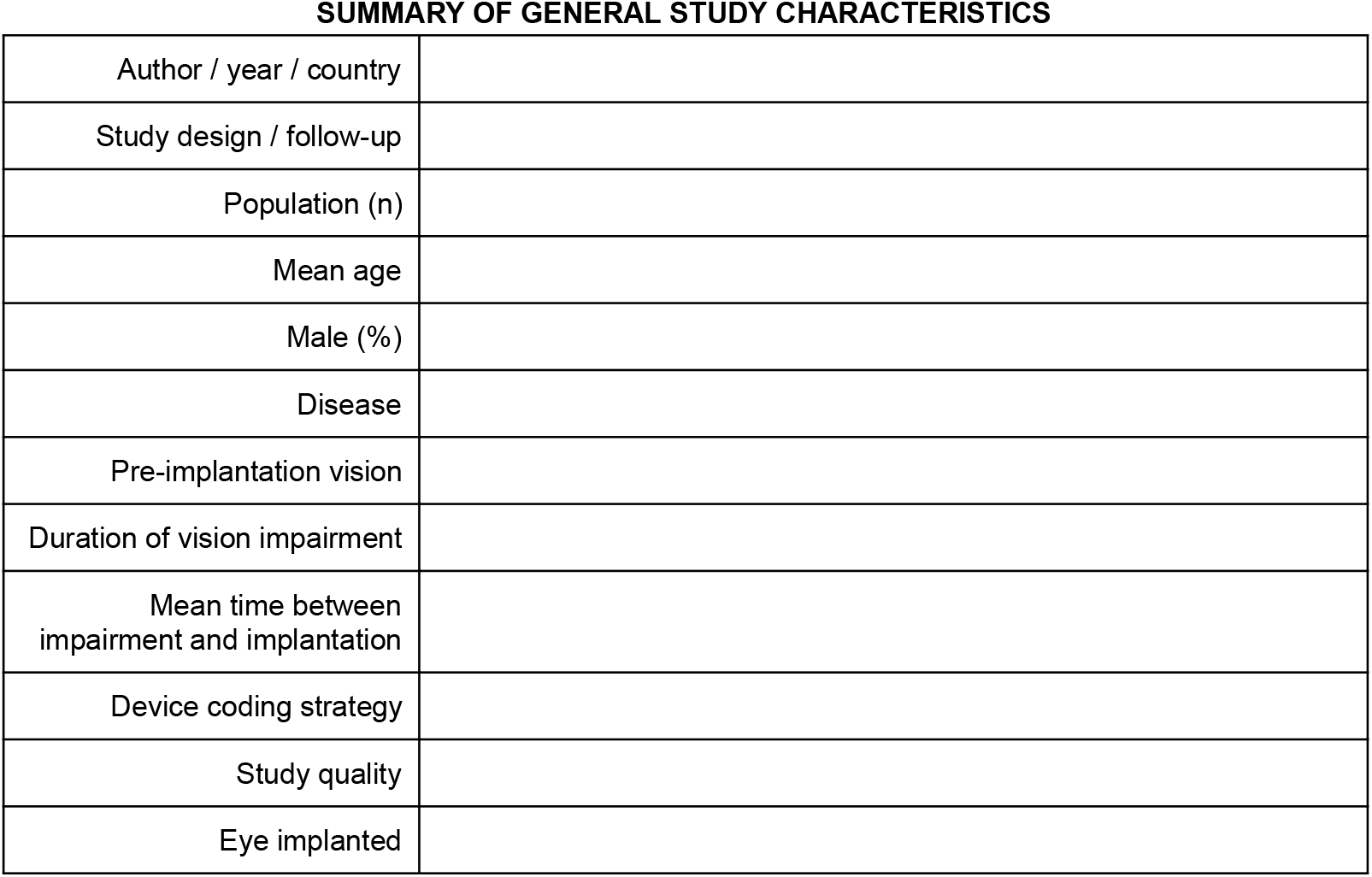
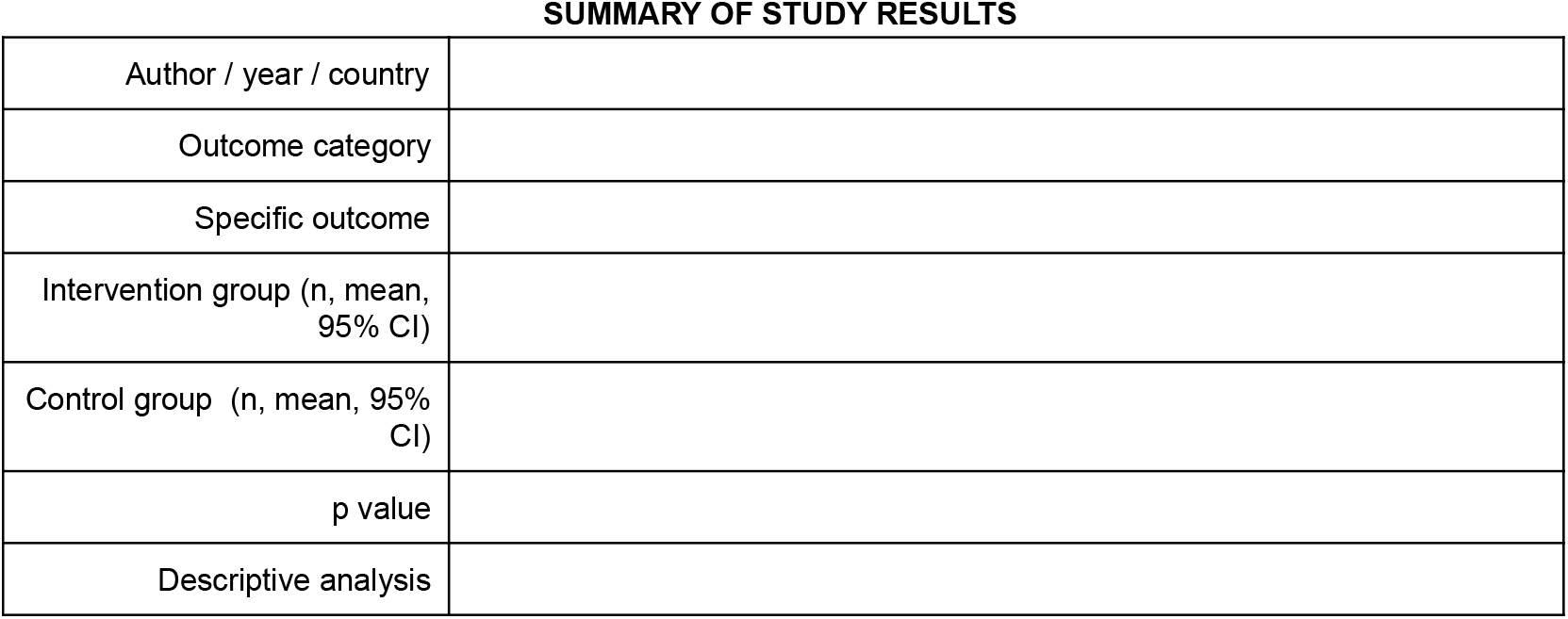
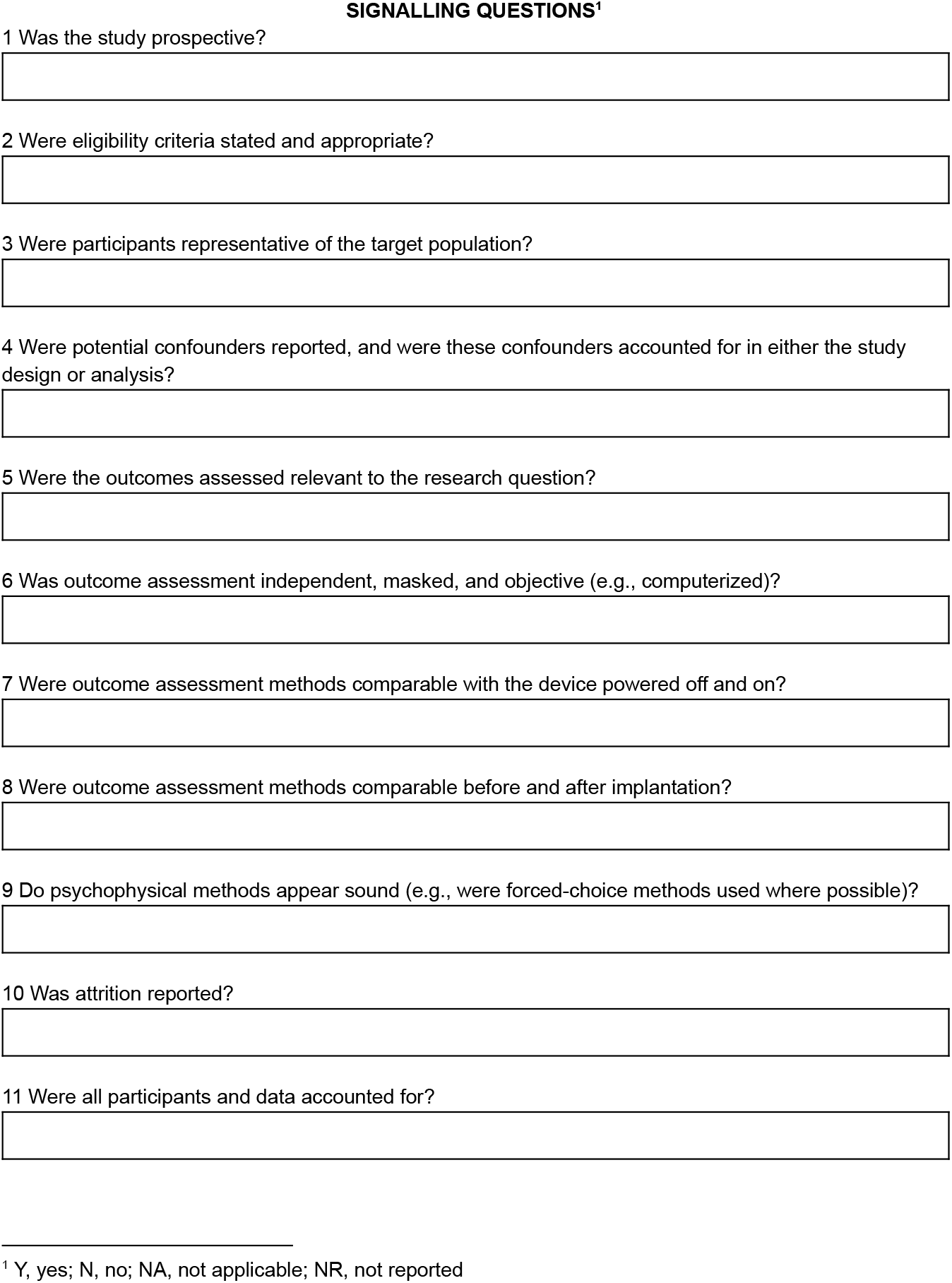
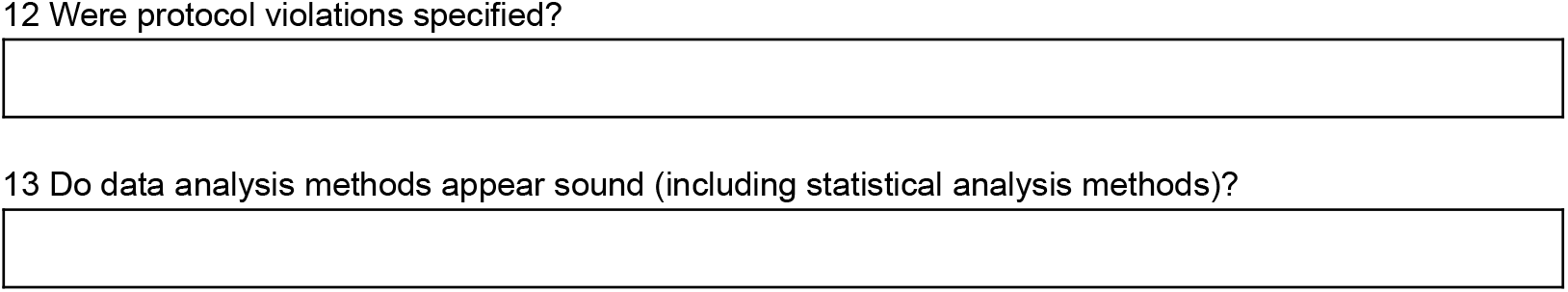

## References

1. Hartong DT, Berson EL, Dryja TP. Retinitis pigmentosa. Lancet. 2006; 368:1795–1809.

2. Hamel C. Retinitis pigmentosa. Orphanet Journal of Rare Diseases. 2006; 1:40.

3. Bunker CH, Berson EL, Bromley WC, Hayes RP, Roderick TH. Prevalence of retinitis pigmentosa in Maine. American Journal of Ophthalmology. 1984; 97:357–365.

4. Grøndahl J. Estimation of prognosis and prevalence of retinitis pigmentosa and Usher syndrome in Norway. Clinical Genetics. 1987; 31:255–264.

5. Stone JL, Barlow WE, Humayun MS, de Juan E, Milam AH. Morphometric analysis of macular photoreceptors and ganglion cells in retinas with retinitis pigmentosa. Archives of Ophthalmology. 1992; 110:1634–1639.

6. Weiland JD, Walston ST, Humayun MS. Electrical stimulation of the retina to produce artificial vision. Annual Review of Vision Science. 2016; 2:273–294.

7. Edwards TL, Cottriall CL, Xue K, Simunovic MP, Ramsden JD, Zrenner E, MacLaren RE. Assessment of the electronic retinal implant alpha AMS in restoring vision to blind patients with end-stage retinitis pigmentosa. Ophthalmology. 2018; 125:432–443.

8. Schaffrath K, Schellhase H, Walter P, et al. One-year safety and performance assessment of the Argus II retinal prosthesis: a post-approval study. JAMA Ophthalmology. 2019; 137:896–902.

9. Fujikado T, Kamei M, Sakaguchi H, et al. One-year outcome of 49-channel suprachoroidal–transretinal stimulation prosthesis in patients with advanced retinitis pigmentosa. Investigative Ophthalmology & Visual Science. 2016; 57:6147–6157.

10. Petoe MA, McCarthy CD, Shivdasani MN, et al. Determining the contribution of retinotopic discrimination to localization performance with a suprachoroidal retinal prosthesis. Investigative Ophthalmology & Visual Science. 2017; 58:3231–3239.

11. Shivdasani MN, Sinclair NC, Dimitrov PN, et al. Factors affecting perceptual thresholds in a suprachoroidal retinal prosthesis. Investigative Ophthalmology & Visual Science. 2014; 55:6467–6481.

12. Gaylor JM, Raman G, Chung M, et al. Cochlear implantation in adults: a systematic review and meta-analysis. JAMA Otolaryngology–Head & Neck Surgery. 2013; 139:265–272.

13. Higgins JPT, Savović J, Page MJ, Elbers RG, Sterne JAC (2019). Chapter 8: Assessing risk of bias in a randomized trial. Draft version (29 January 2019) for inclusion in: Higgins JPT, Thomas J, Chandler J, Cumpston M, Li T, Page MJ, Welch VA (editors). Cochrane Handbook for Systematic Reviews of Interventions. Cochrane.

14. Sterne JAC, Hernán MA, McAleenan A, Reeves BC, Higgins JPT. Chapter 25: Assessing risk of bias in a non-randomized study. Draft version (29 January 2019) for inclusion in: Higgins JPT, Thomas J, Chandler J, Cumpston M, Li T, Page MJ, Welch VA (editors). Cochrane Handbook for Systematic Reviews of Interventions. 2019. Cochrane.

15. Moher D, Liberati A, Tetzlaff J, Altman DG, & the PRISMA Group. Preferred Reporting Items for Systematic Reviews and Meta-Analyses: The PRISMA Statement. Annals of Internal Medicine. 2009; 151:264–269.

16. Bach M, Wilke M, Wilhelm B, Zrenner E, Wilke R. Basic quantitative assessment of visual performance in patients with very low vision. Investigative Ophthalmology & Visual Science. 2010; 51:1255–1260.

17. Barnes N, Scott AF, Lieby P, et al. Vision function testing for a suprachoroidal retinal prosthesis: effects of image filtering. Journal of Neural Engineering. 2016; 13:036013.

18. Ahuja AK, Dorn JD, Caspi A, et al. Blind subjects implanted with the Argus II retinal prosthesis are able to improve performance in a spatial-motor task. British Journal of Ophthalmology. 2011; 95:539–543.

19. da Cruz L, Dorn JD, Humayun MS, et al. Five-year safety and performance results from the Argus II retinal prosthesis system clinical trial. Ophthalmology. 2016; 123:2248–2254.

20. Luo YHL, Zhong JJ, da Cruz L. The use of Argus II retinal prosthesis by blind subjects to achieve localisation and prehension of objects in 3-dimensional space. Graefe’s Archive for Clinical and Experimental Ophthalmology. 2015; 253:1907–1914.

21. He Y, Huang NT, Caspi A, Roy A, Montezuma SR. Trade-off between field-of-view and resolution in the thermal-integrated Argus II system. Translational Vision Science & Technology. 2019; 8:29.

22. Hafed ZM, Stingl K, Bartz-Schmidt KU, Gekeler F, Zrenner E. Oculomotor behavior of blind patients seeing with a subretinal visual implant. Vision Research. 2016; 118:119–131.

23. Castaldi E, Cicchini GM, Cinelli L, Biagi L, Rizzo S, Morrone MC. Visual BOLD response in late blind subjects with Argus II retinal prosthesis. PLoS Biology. 2016; 14:e1002569.

24. Parmeggiani F, De Nadai K, Piovan A, Binotto A, Zamengo S, Chizzolini M. Optical coherence tomography imaging in the management of the Argus II retinal prosthesis system. European Journal of Ophthalmology. 2107; 27:16–21.

25. Endo T, Fujikado T, Hirota M, Kanda H, Morimoto T, Nishida K. Light localization with low-contrast targets in a patient implanted with a suprachoroidal–transretinal stimulation retinal prosthesis. Graefe’s Archive for Clinical and Experimental Ophthalmology. 2018; 256:1723–1729.

26. Barry MP, Dagnelie G. Hand-camera coordination varies over time in users of the Argus II retinal prosthesis system. Frontiers in Systems Neuroscience. 2016; 10:41.

27. Caspi A, Roy A, Wuyyuru V, et al. Eye movement control in the Argus II retinal-prosthesis enables reduced head movement and better localization precision. Investigative Ophthalmology & Visual Science. 2018; 59:792–802.

28. Hsu PC, Chen PY, Chung YS, Lin TC, Hwang DK, Chen SJ, Kao CL. First implantation of retinal prosthesis in a patient with high myopia after surgery and rehabilitation program in Taiwan. Journal of the Chinese Medical Association. 2019; 82:599–602.

29. Ho AC, Humayun MS, Dorn JD, et al. Long-term results from an epiretinal prosthesis to restore sight to the blind. Ophthalmology. 2015; 122:1547–1554.

30. Stingl K, Bartz-Schmidt KU, Besch D, et al. Subretinal visual implant alpha IMS–clinical trial interim report. Vision Research. 2015; 111:149–160.

31. Stingl K, Schippert R, Bartz-Schmidt KU, et al. Interim results of a multicenter trial with the new electronic subretinal implant Alpha AMS in 15 patients blind from inherited retinal degenerations. Frontiers in Neuroscience. 2017; 11:445.

32. Takhchidi KhP, Takhchidi NKh, Manoyan RA, Gliznitsa PV. A bionic eye: performance of the Argus II retinal prosthesis in low-vision and social rehabilitation of patients with end-stage retinitis pigmentosa. Bulletin of the Russian State Medical University. 2019; 3:52–57.

33. Yue L, Falabella P, Christopher P, et al. Ten-year follow-up of a blind patient chronically implanted with epiretinal prosthesis Argus I. Ophthalmology. 2015; 122:2545–2552.

34. Grant DA, Hake HW, Hornseth JP. Acquisition and extinction of a verbal conditioned response with differing percentages of reinforcement. Journal of Experimental Psychology. 1951; 42:1.

35. Wozny DR, Beierholm UR, Shams L. Probability matching as a computational strategy used in perception. PLoS Computational Biology. 2010; 6:e1000871.

36. Dorn JD, Ahuja AK, Caspi A, et al. The detection of motion by blind subjects with the epiretinal 60-electrode (Argus II) retinal prosthesis. JAMA Ophthalmology. 2013; 131:183–189.

37. Garcia S, Petrini K, Rubin GS, da Cruz L, Nardini M. Visual and non-visual navigation in blind patients with a retinal prosthesis. PLoS ONE. 2015; 10:e0134369.

38. Dagnelie G, Christopher P, Arditi A, et al. Performance of real-world functional vision tasks by blind subjects improves after implantation with the Argus II retinal prosthesis system. Clinical & Experimental Ophthalmology. 2017; 45:152–159.

39. Muqit MM, Velikay-Parel M, Weber M, et al. Six-Month Safety and Efficacy of the Intelligent Retinal Implant System II Device in Retinitis Pigmentosa. Ophthalmology. 2019; 126:637–639.

40. Geruschat DR, Richards TP, Arditi A, et al. An analysis of observer-rated functional vision in patients implanted with the Argus II Retinal Prosthesis System at three years. Clinical and Experimental Optometry. 2016; 99:227–232.

41. Duncan JL, Richards TP, Arditi A, et al. Improvements in vision-related quality of life in blind patients implanted with the Argus II Epiretinal Prosthesis. Clinical and Experimental Optometry. 2017; 100:144–150.

42. Graham NVS. Visual Pattern Analyzers. 1989. Oxford University Press.

43. Tyler RS, Abbas P, Tye-Murray N, et al. Evaluation of five different cochlear implant designs: audiologic assessment and predictors of performance. The Laryngoscope. 1988; 98:1100–1106.

44. Karanicolas PJ, Farrokhyar F, Bhandari M. Blinding: Who, what, when, why, how? Canadian Journal of Surgery. 2010; 53:345.

45. Schulz KF, Grimes DA. Blinding in randomised trials: hiding who got what. Lancet. 2002; 359:696–700.

46. Wood L, Egger M, Gluud LL, et al. Empirical evidence of bias in treatment effect estimates in controlled trials with different interventions and outcomes: meta-epidemiological study. British Medical Journal. 2008; 336:601–605.

47. Porta M (Ed.). A Dictionary of Epidemiology. 2014. Oxford University Press.

48. Kaptchuk TJ, Goldman P, Stone DA, Stason WB. Do medical devices have enhanced placebo effects? Journal of Clinical Epidemiology. 2000; 53:786–792.

49. Bach M. The “Freiburg visual acuity test” - automatic measurement of visual acuity. Optometry and Vision Science. 1996; 73:49–63.

50. Stingl K, Bartz-Schmidt KU, Besch D, et al. Artificial vision with wirelessly powered subretinal electronic implant alpha-IMS. Proceedings of the Royal Society B: Biological Sciences. 2013; 280:20130077.

51. Yanai D, Weiland JD, Mahadevappa M, Greenberg RJ, Fine I, Humayun MS. Visual performance using a retinal prosthesis in three subjects with retinitis pigmentosa. American Journal of Ophthalmology. 2007; 143:820–827.

52. Luo YHL, da Cruz L. The Argus II retinal prosthesis system. Progress in Retinal and Eye Research. 2016; 50:89–107.

53. Rizzo S, Cinelli L, Finocchio L, Tartaro R, Santoro F, Gregori NZ. Assessment of Postoperative Morphologic Retinal Changes by Optical Coherence Tomography in Recipients of an Electronic Retinal Prosthesis Implant. JAMA Ophthalmology. 2019; 137:272–278.

54. Demchinsky AM, Shaimov TB, Goranskaya DN, Moiseeva IV, Kuznetsov DI, Kuleshov DS, Polikanov DV. The first deaf-blind patient in Russia with Argus II retinal prosthesis system: what he sees and why. Journal of Neural Engineering. 2019; 16:025002.

55. Humayun MS, de Juan Jr E, Dagnelie G. The bionic eye: a quarter century of retinal prosthesis research and development. Ophthalmology. 2016; 123:S89–S97.

56. Ayton LN, Rizzo JF, Bailey IL, et al. Harmonization of outcomes and vision endpoints in vision restoration trials: recommendations from the international HOVER taskforce. Translational Vision Science & Technology. 2020; 9: 25.

57. Hallum LE, Suaning GJ, Taubman DS, Lovell NH. Simulated prosthetic visual fixation, saccade, and smooth pursuit. Vision Research. 2005; 45: 775–788.

58. Chen SC, Hallum LE, Suaning GJ, Lovell NH. A quantitative analysis of head movement behaviour during visual acuity assessment under prosthetic vision simulation. Journal of Neural Engineering. 2007; 4: S108.

59. Peli E. Testing vision is not testing for vision. Translational Vision Science & Technology. 2020; 9(13): 32.

60. Adeyemo O, Jeter PE, Rozanski C, et al. Living with ultra-low vision: an inventory of self-reported visually guided activities by individuals with profound visual impairment. Translational Vision Science & Technology. 2017;6(3): 10.

